# Using sero-epidemiology to monitor disparities in vaccination and infection with SARS-CoV-2

**DOI:** 10.1101/2021.10.06.21264573

**Authors:** Isobel Routledge, Saki Takahashi, Adrienne Epstein, Jill Hakim, Owen Janson, Keirstinne Turcios, Jo Vinden, John Tomas Risos, Margaret Rose Baniqued, Lori Pham, Clara Di Germanio, Michael Busch, Margot Kushel, Bryan Greenhouse, Isabel Rodríguez-Barraquer

## Abstract

**Background:** As COVID-19 vaccines continue to be rolled-out, the “double burden” of health disparities in both exposure to infection and vaccination coverage intersect to determine the current and future patterns of infection, immunity, and mortality. Serology provides a unique opportunity to measure biomarkers of infection and vaccination simultaneously, and to relate these metrics to demographic and geographic factors.

**Methods:** Leveraging algorithmically selected residual serum samples from two hospital networks in San Francisco, we sampled 1014 individuals during February 2021, capturing transmission during the first 11 months of the epidemic and the early roll out of vaccination. These samples were tested using two serologic assays: one detecting antibodies elicited by infection, and not by vaccines, and one detecting antibodies elicited by both infection and vaccination. We used Bayesian statistical models to estimate the proportion of the population that was naturally infected and the proportion protected due to vaccination.

**Findings:** We estimated that the risk of prior infection of Latinx residents was 5.3 (95% CI: 3.2 - 10.3) times greater than the risk of white residents aged 18-64 and that white San Francisco residents over the age of 65 were twice as likely (2.0, 95% CI: 1.1 - 4.6) to be vaccinated as Black residents. We also found socioeconomically deprived zipcodes in the city had high probabilities of natural infections and lower vaccination coverage than wealthier zipcodes.

**Interpretation:** Using a platform we created for SARS-CoV-2 serologic data collection in San Francisco, we characterized and quantified the stark disparities in infection rates and vaccine coverage by demographic groups over the first year of the pandemic. While the arrival of the SARS-CoV-2 vaccine has created a ‘light at the end of the tunnel’ for this pandemic, ongoing challenges in achieving and maintaining equity must also be considered.

**Funding:** NIH, NIGMS, Schmidt Science Fellows in partnership with the Rhodes Trust and the Chan Zuckerberg Biohub.

## Background

During the initial waves of the COVID-19 pandemic, disparities in disease burden were largely driven by differences in infection rates, as a result of factors including occupation, ability to shelter in place or to take sick leave, access to testing, housing status and crowding, and neighborhood exposure. In addition to driving disparities in infection rates with this novel virus, existing structural inequalities are associated with disparities in the risk of comorbidities such as diabetes and heart disease (as a result of factors such as, but not limited to, nutrition, access to exercise and increased stress), which increase the likelihood of hospitalization and death from COVID-19, and with disparities in access to healthcare both in managing comorbidities and in accessing care for COVID-19.

As vaccine roll-outs advance in the United States and globally, there are disparities in both vaccine access and uptake. These disparities are multifactorial and complex, including reduced technology access and literacy^1^, barriers in access to healthcare, concern about the safety of the vaccines^2^, mistrust as a result of a history of medical racism and discrimination, and poor access to reliable information about the vaccine. In the age of vaccination, policymakers must understand the way in which societal structures affect disparities in both infection and vaccination. These disparities may interact to affect both population level immunity and the burden of COVID-19 in different communities. This is relevant both in the present and in the future, as policymakers consider the continued roll-out of vaccines in the context of new variants, as well as preparing for and responding to other diseases.

Given the high levels of disease under-ascertainment, serology (i.e., the measurement of antibodies) has been particularly useful for understanding SARS-CoV-2 infection levels in the population. When there is variability in testing rates and healthcare seeking behavior, serology is an even more useful tool. Serology provides a unique opportunity to measure biomarkers of infection and vaccination simultaneously, and to relate these metrics to demographic and geographic factors. In settings where vaccines based on the SARS-CoV-2 spike protein (e.g., currently available mRNA or adenovirus vector vaccines) are used, measuring long-lived antibody responses to both spike and non-spike proteins can be used to disentangle immune responses elicited by infection from vaccination. While structural inequalities are by no means limited to the United States, here we focus on a case example leveraging serology to understand inequalities from a domestic perspective.

Even in San Francisco, a city which has had a relatively successful early and sustained COVID-19 response, and has achieved high vaccination coverage over the past few months, reported case counts of COVID-19 and hospitalization rates have been higher in socioeconomically deprived areas, amongst homeless individuals, and within the city’s Latinx and Black communities^3,4^. Disparities in vaccination coverage have also been reported, particularly in the early months of vaccine roll-out, creating a double burden for some vulnerable communities. In San Francisco, whilst some disparities have now been addressed, vaccination remains much lower in homeless individuals and in Black/African American individuals^5^.

To measure disparities in both infection rates and vaccination, we leveraged a SARS-CoV-2 serosurveillance platform launched in March 2020 that utilizes residual blood samples taken from two hospital networks in San Francisco. Estimates derived from this platform during the first wave of the pandemic showed seroprevalence in Latinx individuals to be nearly two times higher than in white individuals, and nearly two times higher in homeless individuals than the population average^6^. We collected samples from 1,014 individuals undergoing routine blood draws between February 4 and February 17, 2021, capturing transmission during the first 11 months of the epidemic and the early roll-out of vaccination for those over 65 years old. These samples were tested using two serologic assays: one detecting antibodies to SARS-CoV-2 elicited by infection and not by vaccines currently used in the US, and one detecting antibodies to SARS-CoV-2 elicited by both infection and vaccination. We used Bayesian statistical models to estimate the proportion of the population that was seropositive due to natural infection and the proportion seropositive due to vaccination, stratified by age, race and ZIP code of residence.

## Methods

### Data

As part of an existing serological survey^6^, residual serum samples from routine blood draws from the University of California, San Francisco (UCSF) and San Francisco Department of Public Health (SFDPH) inpatient and outpatient healthcare systems were sampled between February 4 and February 17, 2021. A total of 1,091 samples were collected, of which 77 were excluded due to participation in a separate COVID research study, and a further 15 were later excluded from further analyses as they could not be linked to antibody test results. The characteristics of the samples are illustrated in Table 1. The full inclusion and exclusion criteria and sampling algorithm are described previously^6^.

**Table 1:** Sample Characteristics. Table showing the sample size and distribution of the sample population by demography and hospital system, compared to the San Francisco Population as determined by the American Community Survey 2019.

|  | N | % | SF Population (ACS 2019) |
| --- | --- | --- | --- |
| **Age** |  |  |  |
| 0-17 | 21 | 2.10% | 13.40% |
| 18-34 | 157 | 15.50% | 30.60% |
| 35-64 | 442 | 43.60% | 40.60% |
| 65+ | 393 | 38.80% | 15.40% |
| Unknown | 1 | 0.10% | N/A |
| **Sex** |  |  |  |
| Female | 509 | 50.20% | 49.30% |
| Male | 504 | 49.70% | 50.70% |
| Unknown | 1 | 0.10% | N/A |
| **Hospital** |  |  |  |
| UCSF | 698 | 68.80% | N/A |
| ZSFG | 316 | 31.20% | N/A |
| **Race/Ethnicity** |  |  |  |
| Asian | 282 | 27.80% | 34.60% |
| Black or African American | 115 | 11.30% | 5.20% |
| Hispanic or Latino | 175 | 17.30% | 15.20% |
| Other | 75 | 7.40% | 5.20% |
| White | 367 | 36.20% | 39.80% |

### Laboratory analysis

Each sample (N = 1014) was tested on two commercial SARS-CoV-2 serologic platforms. The Ortho Clinical Diagnostics VITROS Anti-SARS-CoV-2 Total assay measures the total Ig antibody response to the S1 subunit of the SARS-CoV-2 spike (S) protein and therefore is expected to yield a positive results after natural infection or vaccination^7^. The Roche Elecsys Anti-SARS-CoV-2 assay measures the total Ig antibody response to the SARS-CoV-2 nucleocapsid (N) protein^8^ and therefore is expected to yield a positive results after natural infection but not after vaccination with vaccines based on the spike protein. In a previous analysis assessing the test performance characteristics of many SARS-CoV-2 serological assays, we found both assays to exhibit high sensitivity over time following infection^9^.

### Univariate data analysis (by assay)

Seropositivity on the Vitros assay indicates whether or not an individual has had any prior immune response to SARS-CoV-2, either through natural infection and/or vaccination. The SARS-CoV-2 mRNA and adenovirus vector vaccines elicit immune responses to only the S protein of the virus. Therefore, in contexts where these vaccines are used exclusively (such as the United States), seropositivity on the Roche assay indicates whether an individual has had a prior immune response to SARS-CoV-2 via infection. Assuming perfect test performance characteristics, the difference between the proportion seropositive on Vitros and the proportion seropositive on Roche indicates the proportion of the population that has been vaccinated and has not been infected.

We used Binomial models in a Bayesian framework to first estimate seropositivity separately by assay. We adjusted for the manufacturer-reported specificity of each assay (100% for Vitros and 99.80% for Roche) and for in-house estimates of the sensitivity of each assay at 2 months post symptom onset among non-hospitalized individuals based on a longitudinal post-infection study^9^, corresponding to 83.8% for Vitros and 90.0% for Roche. We note that sensitivity was particularly consistent over time following infection for these assays, so our results are robust to the choice of exact time point used. We computed 95% credible intervals (CrI) to quantify uncertainty in posterior estimates. For these univariate analyses, age was stratified into 4 groups (0-17 y, 18-34 y, 35-64 y, and 65+ y) and race/ethnicity was stratified into 5 groups (Asian, Black, Latinx, White, and Other).

### Bivariate data analysis (infection vs. vaccination)

We then conducted bivariate analyses using the results of each sample on both assays. We used the data set of individuals who had a test result on both the Vitros and Roche assays, and removed the 81 samples that had a result on only one assay. In addition, we removed the 3 samples that tested negative on Vitros and positive on Roche, which likely reflects a false negative result on the Vitros assay and/or a false positive result on the Roche assay (Supplementary Table 1).

#### For demographic analyses

Age was stratified into 2 groups (18-64 y and 65+ y), and race/ethnicity was stratified into 4 groups (Asian, Black, Latinx, and White). We omitted individuals aged 0-17 y from this portion of the analysis, as individuals in that age group were not eligible for vaccination during this time frame in San Francisco (February 2021). The decision to choose 65 y as a cutoff was due to the age-based roll-out of SARS-CoV-2 vaccination and the expected resultant differences in vaccine coverage by age. We also omitted individuals with race/ethnicity of “Other” due to lack of data on vaccine doses in this demographic group that are used as a prior for estimation (see below). For each combination of age group and race/ethnicity *j*, we estimated the marginal probabilities of natural infection, Pr(inf), and of vaccination, Pr(vacc), separately as follows.

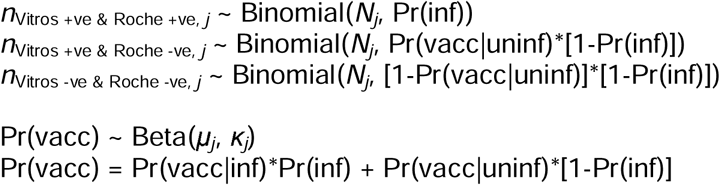

*N*_*j*_ represents the number of individuals in our data set in group *j* who were included, and *n*_*x,j*_ represents the number of individuals in group *j* with antibody results *x*. For this analysis, we assumed perfect test performance characteristics. As Pr(vacc|inf) is not identified by our data, and we do not assume that Pr(vacc|inf) = Pr(vacc|uninf), we set up a process to estimate the hyper-priors μ*j* (mean) and κ*j* (precision) for each group *j* based on reported vaccination coverage data. To do this, we first obtained the total population size *M*_*j*_ of group *j* in San Francisco as well as the reported number of individuals in that group *m*_*j*_ who had been vaccinated up to January 20, 2021 (i.e., 3 weeks before the weighted mean date of sample collection, allowing for time to sero-conversion after vaccination). We then used a hypergeometric distribution to sample *N*_*j*_ individuals without replacement from a population in which *m*_*j*_ individuals had been vaccinated and *M*_*j*_ - *m*_*j*_ had not. We then calculated the empirical proportion of vaccinated individuals among the *N*_*j*_ in that simulation, and repeated this procedure 10,000 times to obtain a prior distribution of Pr(vacc). This distribution was used to estimate the hyper-priors of the Beta distribution. This procedure was performed separately for each group *j*.

For geographic analyses, ZIP codes with fewer than 10 individuals were excluded. As data on vaccine doses distributed by ZIP code during this time-frame was not available, we modified the model above by assuming that vaccination was independent of infection, and estimated a single Pr(inf) and Pr(vacc) for each ZIP code. To measure disparities in both infection rates and vaccination, we leveraged a SARS-CoV-2 serosurveillance platform launched in March 2020 that utilizes residual blood samples taken from two hospital networks in San Francisco. Estimates derived from this platform during the first wave of the pandemic showed seroprevalence in Latinx individuals to be nearly two times higher than in white individuals, and nearly two times higher in homeless individuals than the population average ^6^. We collected samples from 1,014 individuals undergoing routine blood draws between February 4 and February 17, 2021, capturing transmission during the first 11 months of the epidemic and the early roll-out of vaccination. These samples were tested using two serologic assays: one detecting antibodies to SARS-CoV-2 elicited by infection and not by vaccines currently used in the US, and one detecting antibodies to SARS-CoV-2 elicited by both infection and vaccination. We used Bayesian statistical models to estimate the proportion of the population that was seropositive due to natural infection and the proportion seropositive due to vaccination, stratified by age, race and ZIP code of residence.

## Results

Between February 4, 2021, and February 17, 2021, we collected samples, from 1014 individual patients, from UCSF Health (*n*□=□ 698 patients) and the San Francisco Department of Public Health (*n*□=□ 316 patients) networks. By design, the geographic distribution of residents matched the proportion of the San Francisco population living in each zip code (**Fig. 2**). Our sample was equally distributed by sex, however over-represented the 65+ age range and underrepresented the 0-34 age range relative to the San Francisco population (**Table 1**). Our results were relatively representative of the San Francisco population by race and ethnicity, although our sample overrepresented those who identified as Black/African American and slightly underrepresented those who identified as Asian.

**Figure 1:**
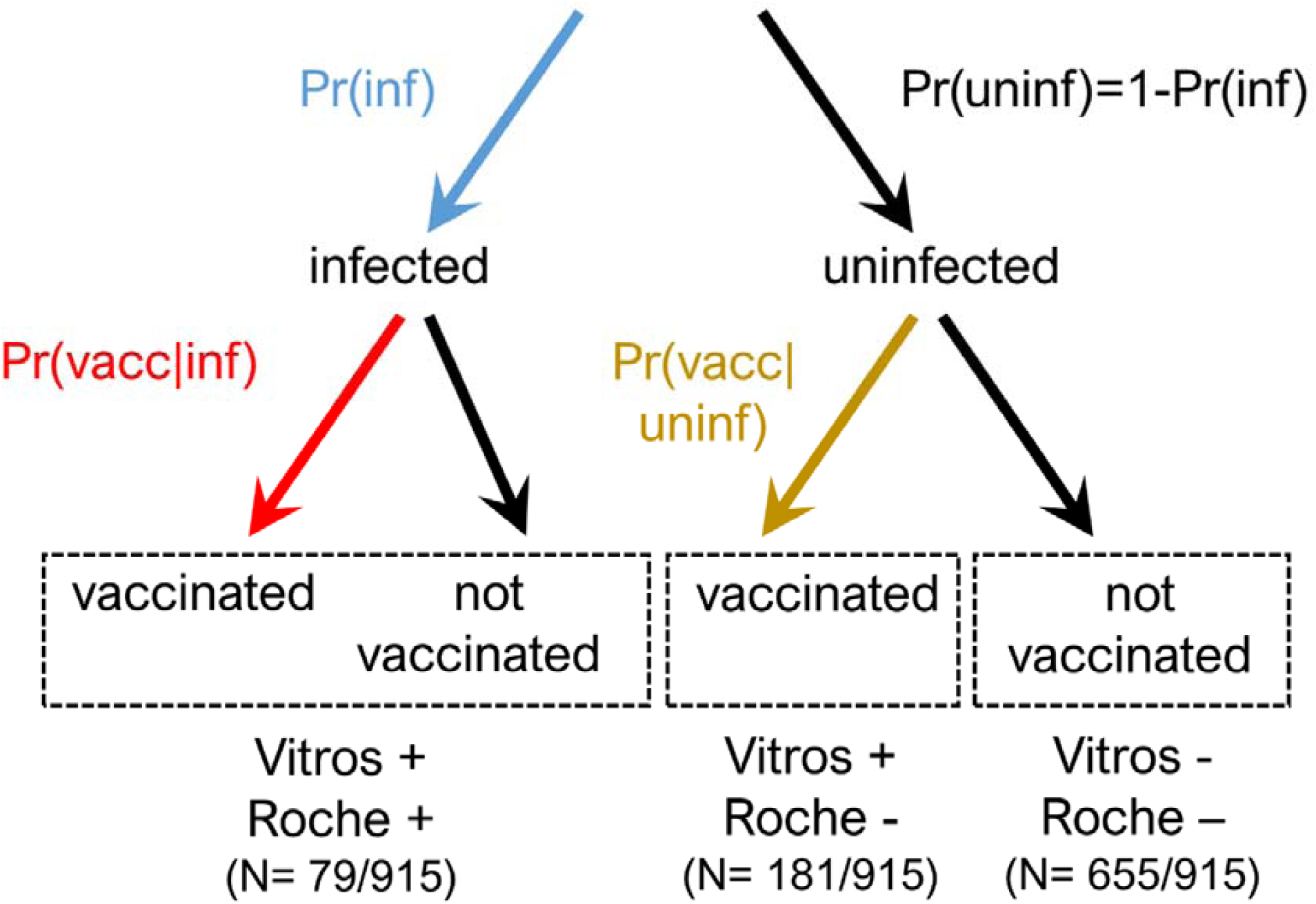
Schematic of parameters to be estimated using serosurveillance platform (shown in red, blue and gold). Red represents the probability of vaccination given prior infection, Pr(vacc|inf), blue represents the probability of prior infection, Pr(inf), and gold represents the probability of vaccination given no prior history of infection, Pr(vacc|uninf).

**Figure 2:**
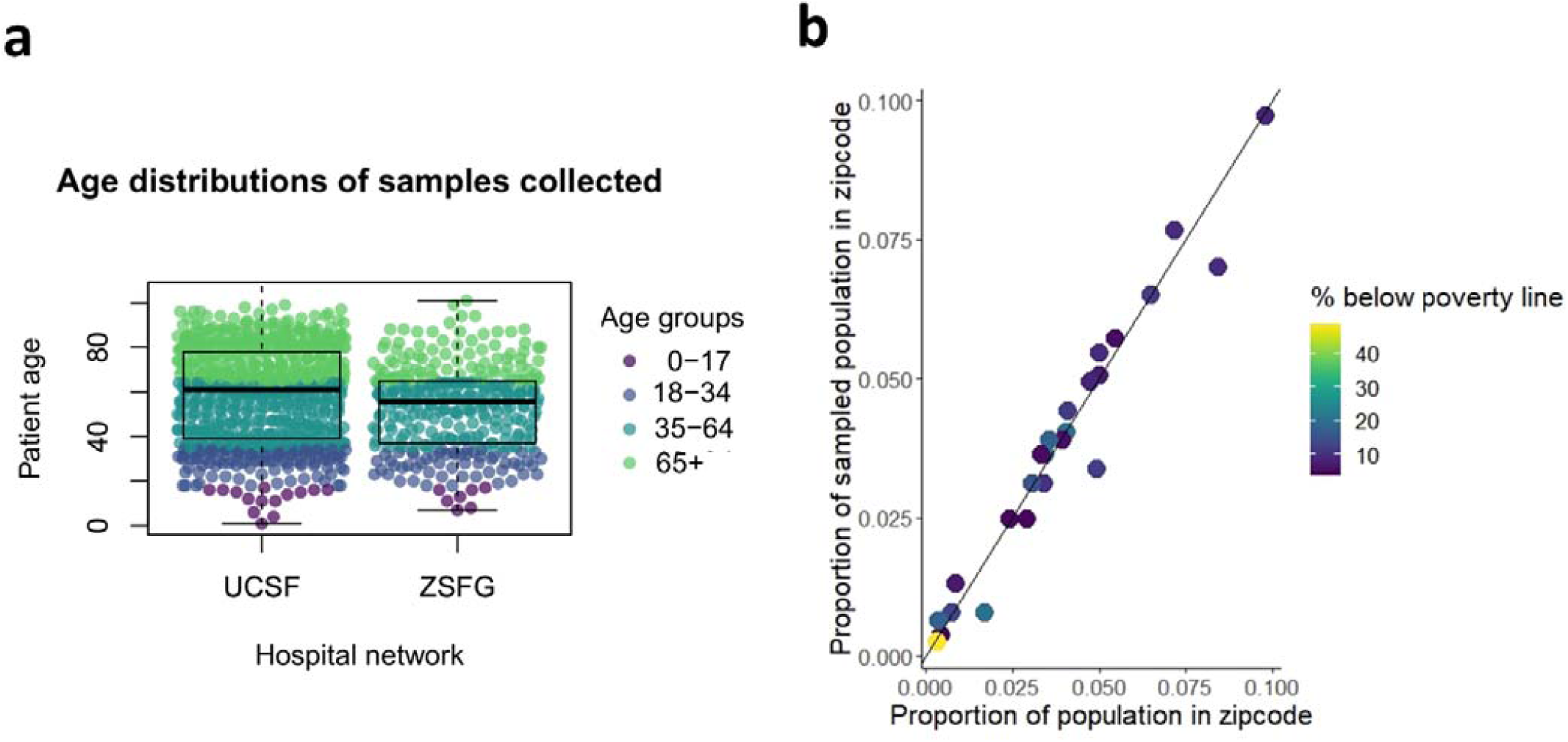
Sample characteristics. **(A)** Age distribution by hospital week of sample collection within the University of California, San Francisco (UCSF) and San Francisco Department of Public Health (ZSFG) hospital networks. Each point represents a sample and colors correspond to age bins used for analysis. **(B)** Proportion of samples from a given San Francisco zipcode plotted against the proportion of the San Francisco population within that zipcode. Colors show the percentage of residents below the poverty line within that zipcode, as determined by the American Community Survey 2019.

Following testing samples on both Vitros and Roche assays, of the sampled population where assay results were complete (N= 915), we found that while 28.4% (N = 260) tested positive on the Vitros assay and therefore antibodies to SARS-CoV-2 were detected, only 8.6% (N = 79) tested positive on the Roche assay, detecting antibodies elicited by prior natural infection (Figure 1). Of the 999 samples where assay results were returned, N= 81 samples were excluded from the analyses due to missing data in at least one assay, and 3 additional samples were excluded due to positive results on the Roche assay despite negative results on the Vitros assay.

Our estimated probabilities of vaccination and prior infection stratified by age, race/ethnicity and ZIP code showed striking differences in prior infection rates and vaccination rates across the city (**Figure 3, Supplementary Table 2**). Across all age and demographic groups, ZIP codes in the southeastern region of the city, comprising medically underserved neighborhoods, had demonstrably higher rates of prior infection and lower rates of vaccination. This pattern is not evident in estimated seroprevalence by the vitros assay which captures antibody responses acquired through natural infection and/or vaccination. For example, within the 94124 zipcode, Bayview-Hunter’s Point, one of the city’s most deprived zipcodes, the mean probability of prior infection was 0.155 (95% CI: 0.077 - 0.254) and vaccination was 0.079 (0.019-0.163), whereas 94115, Pacific Heights, one of the wealthier zipcodes in San Francisco with a median household income almost double that of Bayview-Hunter’s Point at 123,037^10^, the probability of infection was just 0.023 (0.001, 0.080) and vaccination was 0.359 (0.258 - 0.467).

**Figure 3:**
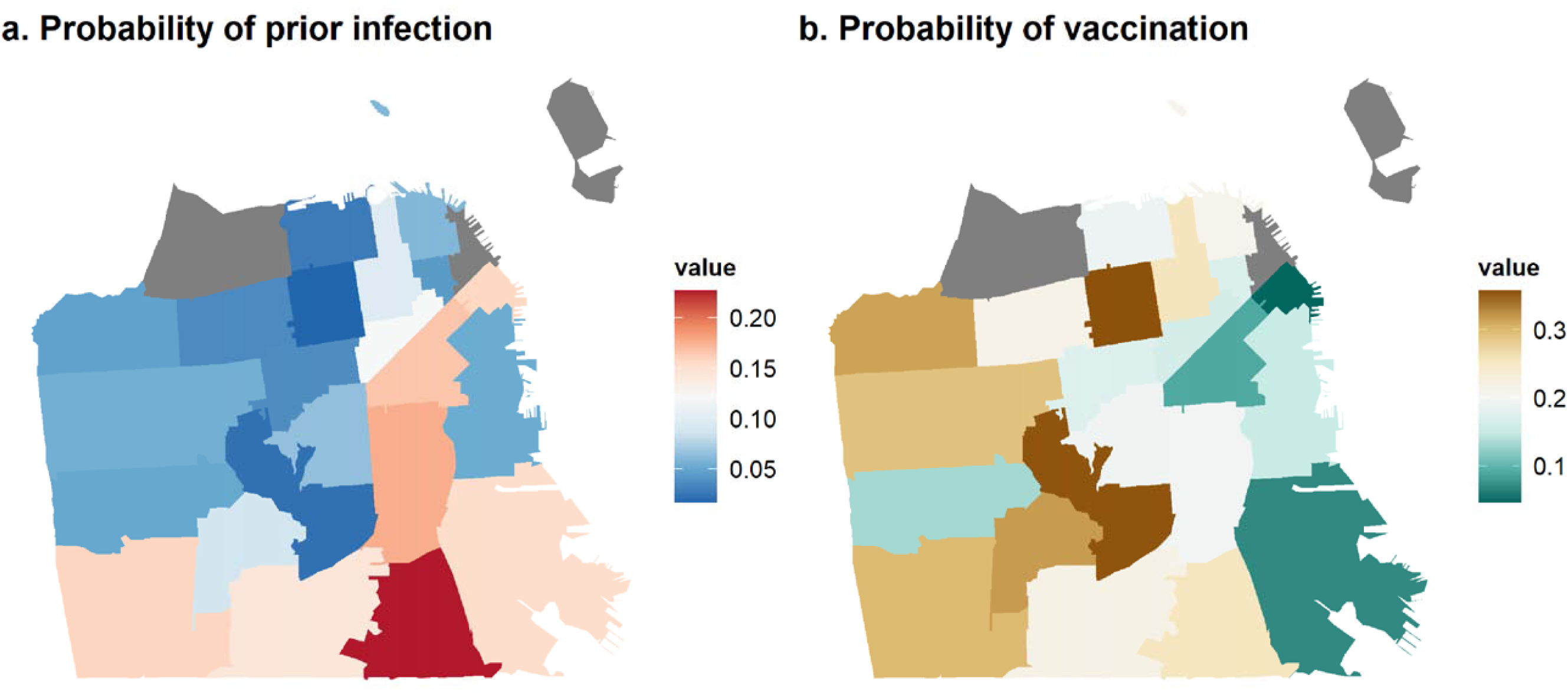
Maps showing geographic disparities in SARS-CoV-2 within San Francisco. Maps show (a) estimated probability of prior infection and (b) probability of vaccination by ZIP code in San Francisco, as of February 2021.

We found the highest seroprevalence as a result of prior infection in younger age groups (using the Roche assay, **Figure 4a, Supplementary Table 3**). We estimated seroprevalence derived from both vaccination and natural infection using the Vitros assay showed much higher seroprevalence in those aged over 65 (**Figure 4b, Supplementary Table 3**), consistent with the eligibility criteria for vaccination in the weeks before the sampling period.

**Figure 4:**
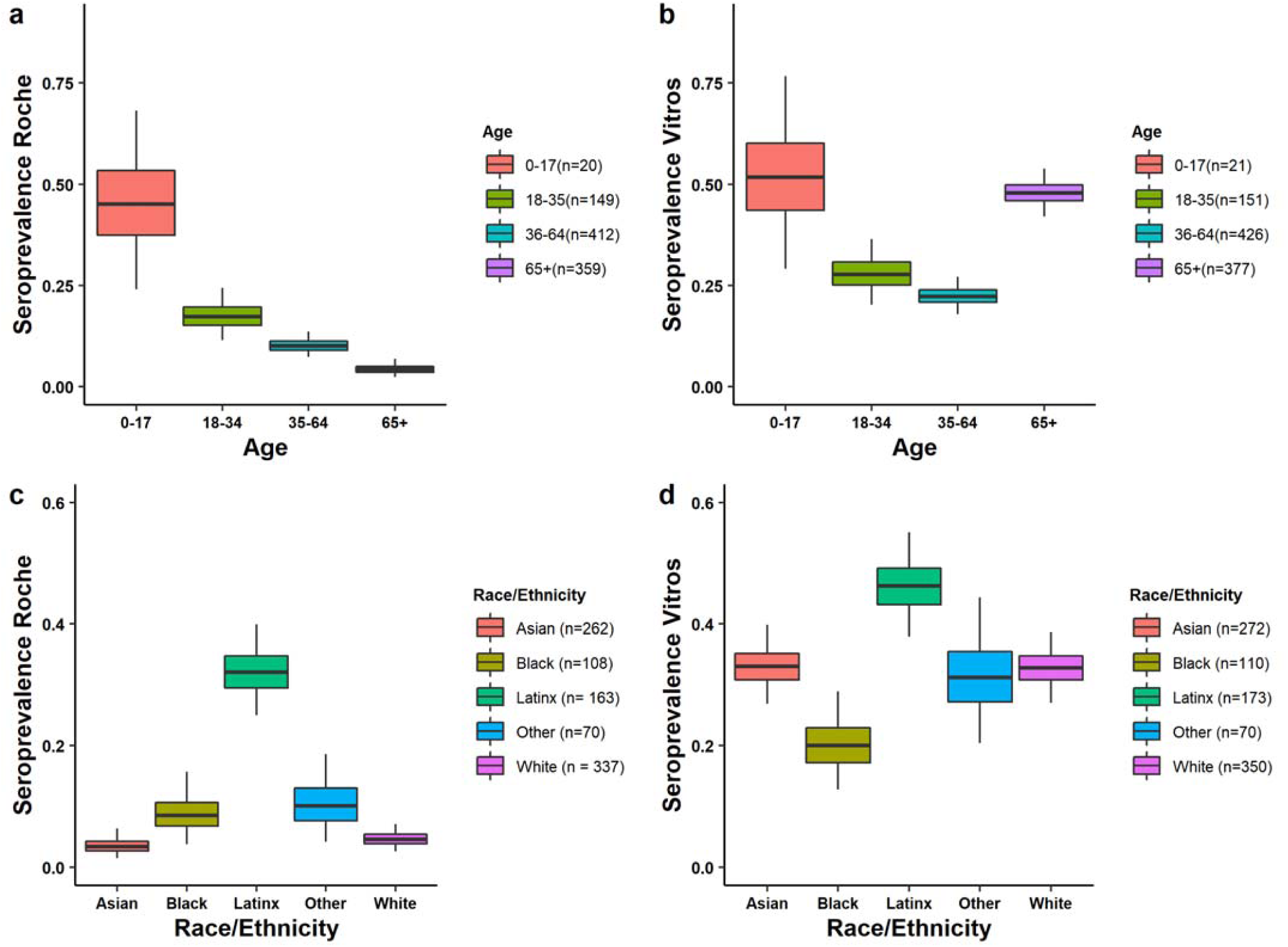
Stratified seroprevalence by assay and by demographic group. **(A)** Univariate Roche seropositivity estimates by age (elicited by prior infection). **(B)** Univariate Vitros estimates by age (elicited by either prior infection or vaccination). **(C)** Univariate Roche estimates by race/ethnicity. **(D)** Univariate Vitros estimates by race/ethnicity.

We identified differences in prior infection rates by race/ethnicity (**Figure 4c-d**): we estimated that the risk of prior infection of Latinx residents was 5.3 (95% CI: 3.2 - 10.3) times greater than the risk of white residents aged 18-64 (**Figure 5a, Supplementary Table 3**). These trends were echoed in older individuals (aged 65+) (**Figure 5a, Supplementary Table 3**). We also identified disparities in vaccination coverage among the 65+ year old population, who were eligible to receive the vaccine during this time period. We estimated that White San Francisco residents over the age of 65 were twice as likely (2.0, 95% CI: 1.1 - 4.6) to be vaccinated as Black residents.

**Figure 5:**
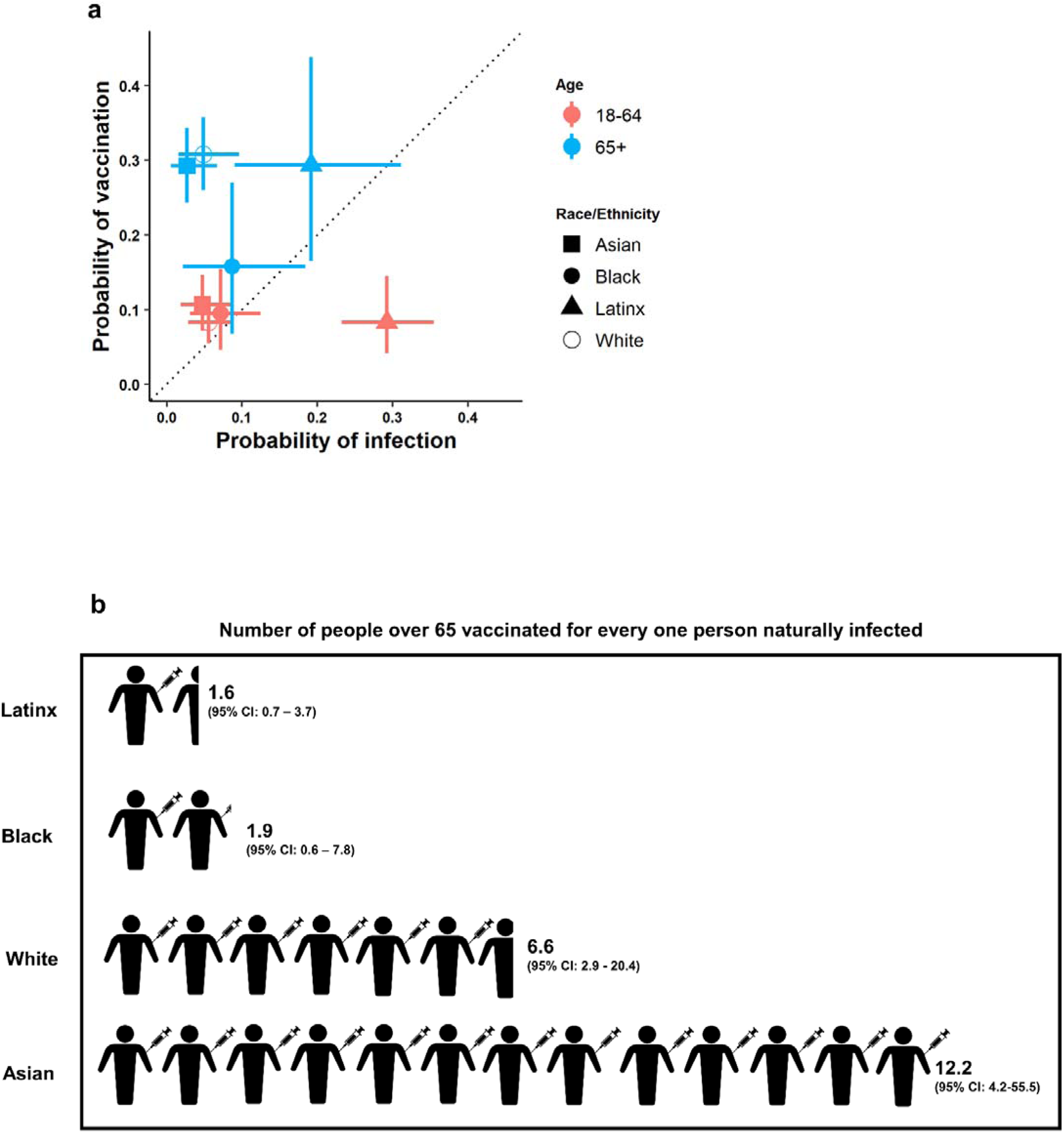
Relationship between probability of vaccination and probability of prior infection by race/ethnicity. **a)** Probability of infection vs. probability of vaccination by age and race/ethnicity. **b)** Infographic showing the number of estimated people vaccinated for every one person previously naturally infected in San Francisco within each racial/demographic group.

Taken together, these findings imply that there is an imbalance between the risk of infection and the rate of vaccination in certain populations.Among the 65+ year old population, we found greatly increased ratios of vaccination compared to infection risk among Asian and white individuals, while this was much lower among Black and Latinx individuals (**Figure 5b, Supplementary Table 4**). For every naturally infected Asian resident of this age group, there were 12.2 vaccinated Asian residents (95%CI: 4.2 - 55.5), whereas for every naturally infected Latinx resident of this age group, there were only 1.6 vaccinated Latinx residents (95% CI: 0.7 – 3.7). For both Latinx and Black individuals over 65 years old, the risk of having immunity acquired through vaccination, relative to natural infection, was up to four times lower than for white individuals.

## Discussion

Using serological data, we quantified disparities in both vaccination coverage and infection rates across different demographic groups and geographies and show that, during early vaccine roll-out, vaccination coverage was much higher in Asian and White populations, despite experiencing lower risk of infection by SARS-CoV-2 than Black and Hispanic/Latinx populations.

The “double burden” we observed in San Francisco during the early vaccine roll-out echoes broader patterns that have been observed in San Francisco and elsewhere. Even though San Francisco was hailed as the first major US city to reach the milestone of 80% vaccination coverage in adults^11^, recent increases in infection have been found to be concentrated in the neighborhoods which were hardest hit by initial infections and where we found vaccination-related immunity was lowest^12^. A report from the University of Texas found striking geographic and racial stratification of cases of COVID-19 and vaccination rates in Austin, Texas, which also closely mapped with indices of deprivation and social vulnerability over ZIP codes^13^. Like in San Francisco, the neighborhoods which were predominantly Latinx communities and had higher indices of deprivation also had higher incidence of SARS-CoV-2 infection and lower vaccination coverage. Disparities in SARS-CoV-2 vaccination coverage among socially vulnerable populations have been documented across the United States^14^ and in other parts of the world^15^.

There are several caveats and limitations to the approach introduced here. The Roche assay can only differentiate antibody responses resulting from natural infections in settings where Spike-based vaccines are used (that do not generate antibody responses against the nucleocapsid), so in geographies where other vaccines are used this approach won’t be suitable. Although we used samples obtained through DPH health network, meaning we included un-insured and under-insured individuals, we still are only able to capture those seeking healthcare or reached by the SF DPH.

While inequalities revealed during COVID-19 are not new, the pandemic has highlighted the ways in which even a city such as San Francisco which invests deeply in public health and social safety nets still has deep structural inequalities, through a combination of higher infection rates, incompatibility of living or work conditions with risk reduction, and lower or delayed access to vaccines as they were rolled out.

Various initiatives are underway around the country to pinpoint geographic and other disparities in the context of COVID-19 (e.g.,^16,17^). However, as well as highlighting disparities, it is important to consider what successful testing and vaccination initiatives may look like and which may be learnt from in future measures. For example, within San Francisco, robust community-academic partnerships have been key for effectively responding to the pandemic in vulnerable communities^18^ and for narrowing gaps in vaccination coverage, such as low-barrier neighborhood vaccination sites^19^. Prospectively, as we gain a better understanding of waning immunity and the potential need for vaccine booster doses, considerations of equity will remain an important consideration for allocating resources. In the context of the United States, where vaccination and infection elicit different immune responses, serology provides a powerful lens through which we can quantify these disparities directly. In addition, it is important to consider the desired metric before conducting a serosurvey, as assays measure different pathways to immunity and any disparities in infection rates may be masked by using assays that measure overall antibody prevalence.

Since the early days of the pandemic, many policy recommendations have been made for ways to reduce health disparities in infection^20^ and vaccination^21^. Policymakers must invest in addressing both the upstream, structural drivers of health disparities, such as providing workers with a living wage, affordable housing, and access to quality healthcare and also downstream drivers such as improved community engagement, targeted testing and vaccination provision, and assistance with common barriers to accessing healthcare such as technology access/literacy, transport, and providing accessible health information in multiple languages. While the arrival of the SARS-CoV-2 vaccine has created a ‘light at the end of the tunnel’ for this pandemic, ongoing challenges that long predate COVID-19 in achieving and maintaining equity must also be considered.

## Data Availability

All code to generate results will be available at https://github.com/EPPIcenter/scale-it-2

## Supplementary Figures and Tables

**Supplementary Figure 1:**
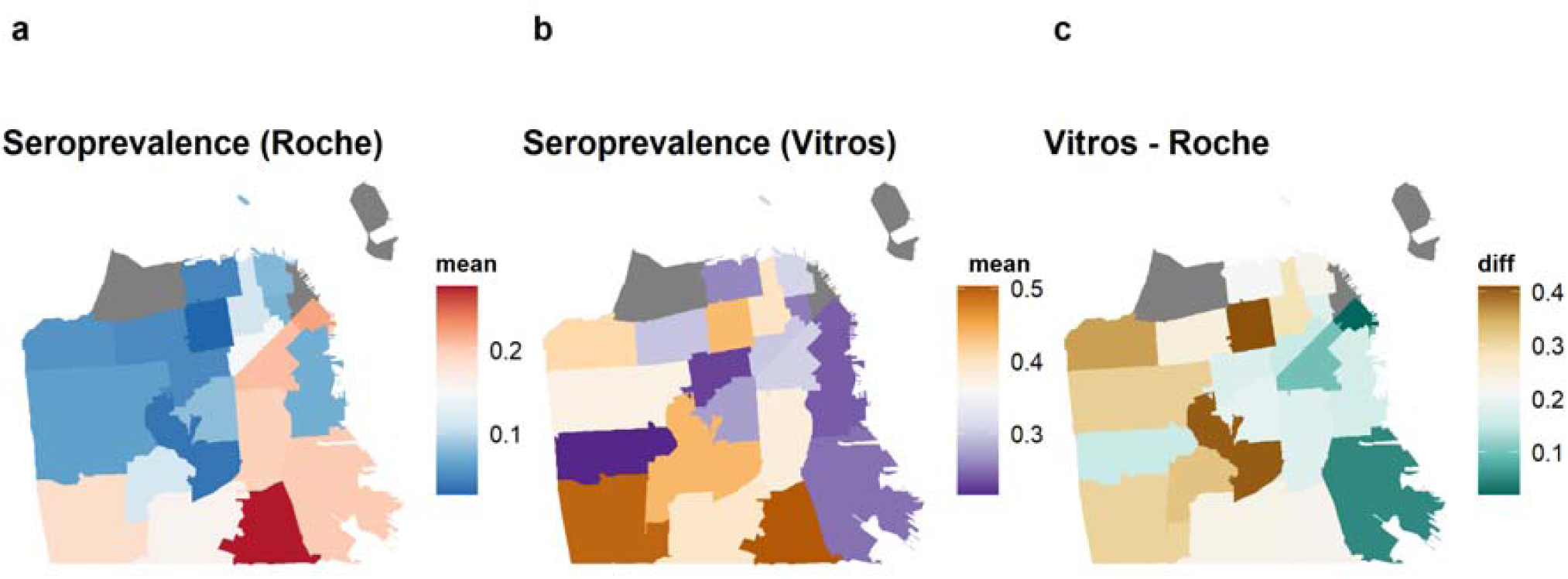
Geographic disparities in SARS-CoV-2 infection and vaccination by ZIP code in San Francisco, as of February 2021 (univariate). **(A)** Adjusted seroprevalence on the Roche assay (reflecting natural infection only). **(B)** Adjusted seroprevalence on the Vitros assay (reflecting both natural infection and/or vaccination). **(C)** The difference in adjusted seroprevalence on the Vitros and Roche assays (reflecting those who were seropositive by vaccination only).

**Supplementary Figure 2:**
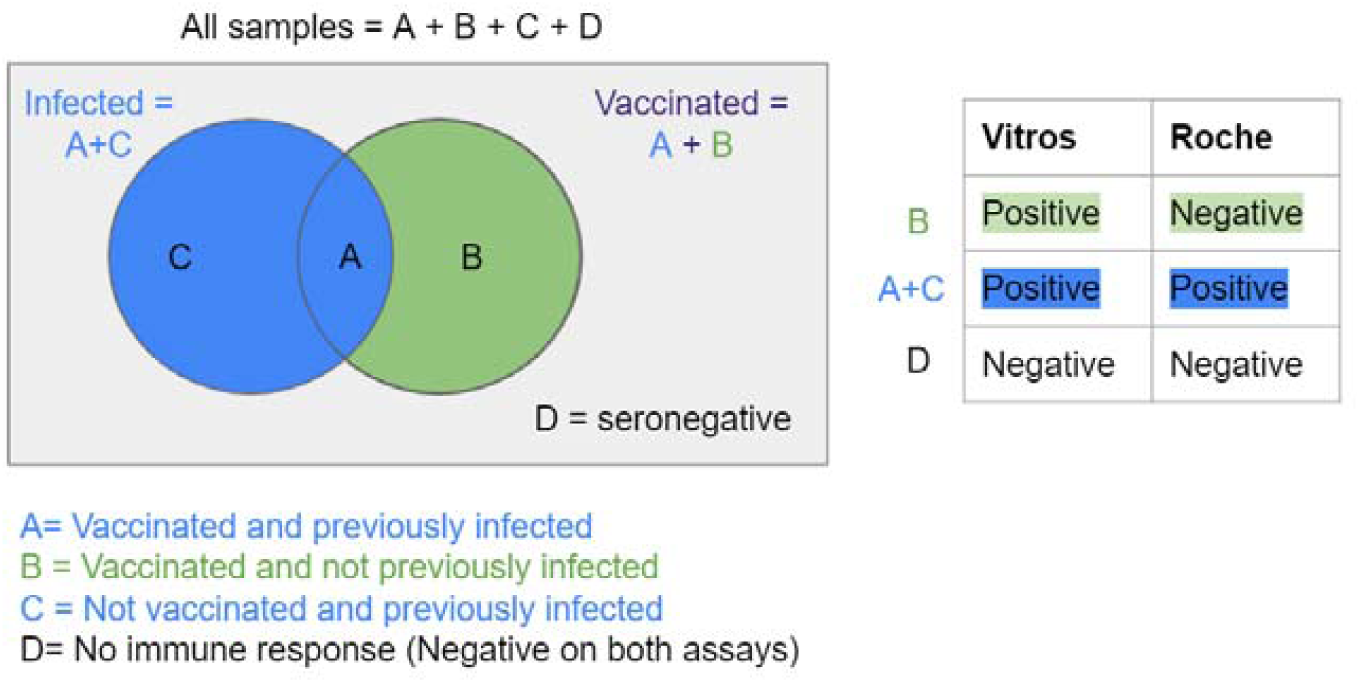
Pathways to immunity via infection and vaccination.

**Supplementary Figure 3:**
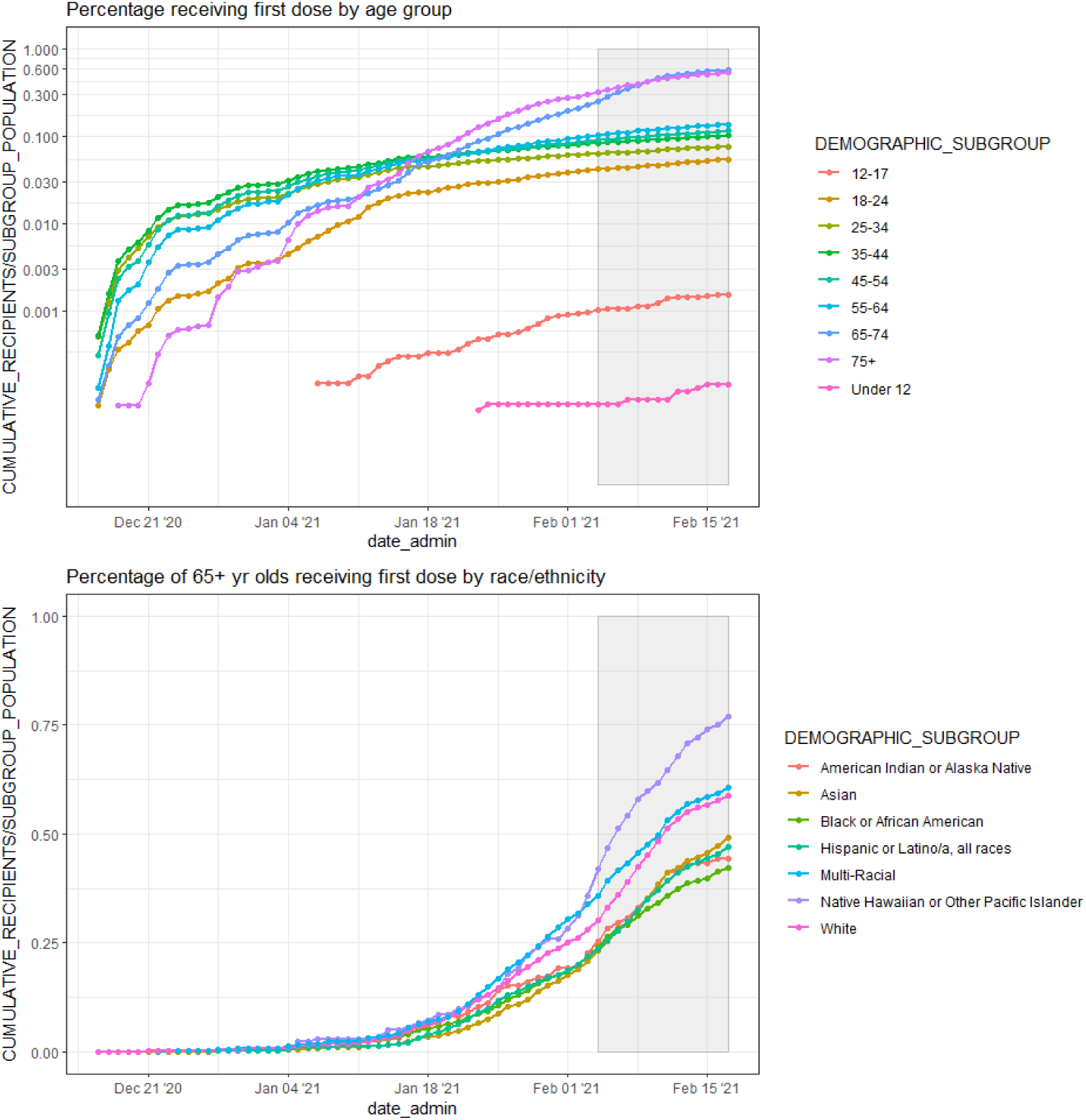
Cumulative proportion of individuals who received the first dose of SARS-CoV-2 vaccine by date, age group, and race/ethnicity. Sampling period of this study shown in grey box. Data downloaded from ^22^.

**Supplementary Figure 4:**
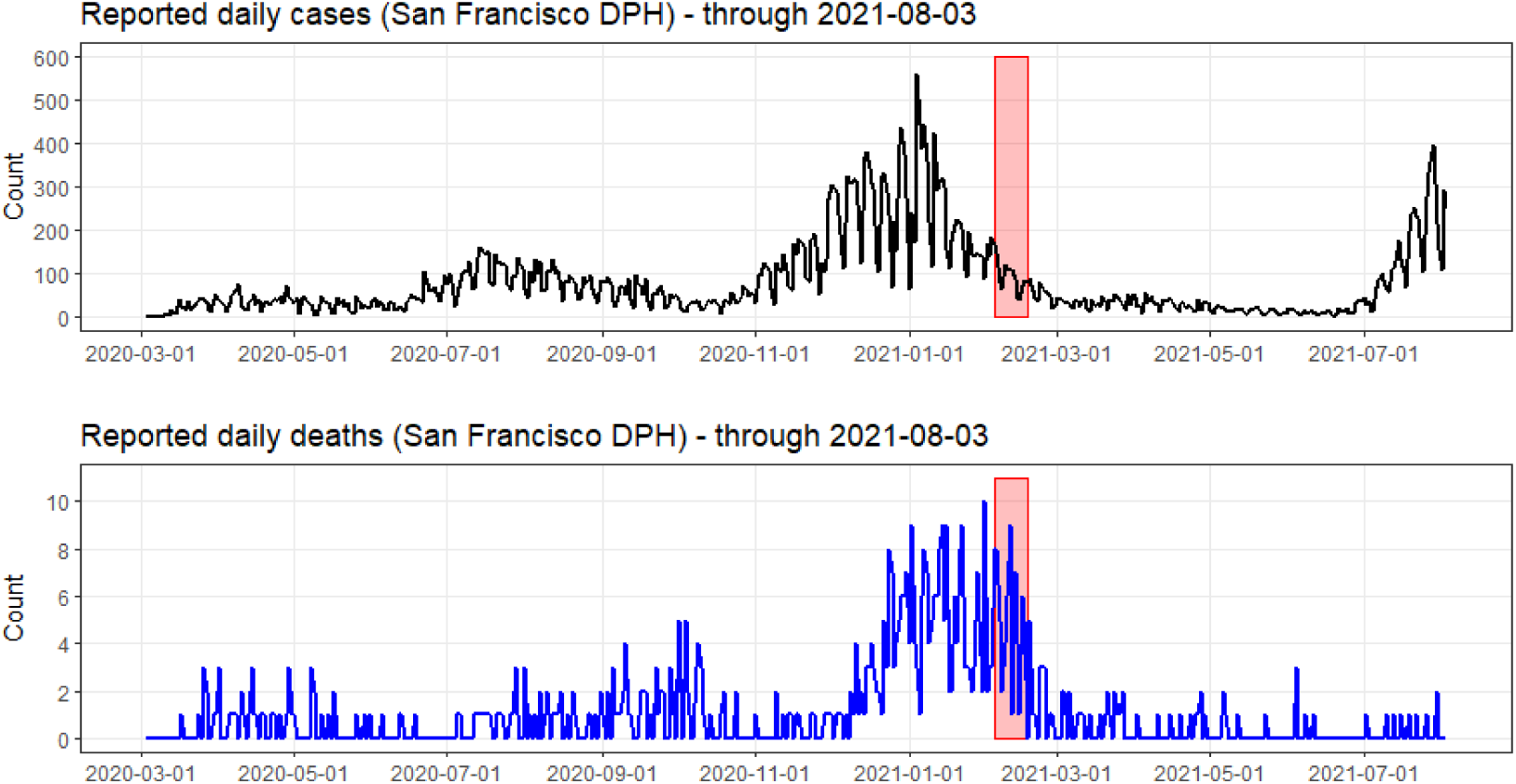
Time series of reported SARS-CoV-2 cases and deaths in San Francisco. Sampling period of this study shown in red box. Data downloaded from ^23^.

**Supplementary Table 1:** Interpretation of bivariate antibody results on the Vitros and Roche assays.

| Vitros result | Roche result | Interpretation for bivariate analyses (n) |
| --- | --- | --- |
| Positive | Positive | Infected and (either vaccinated or unvaccinated) (n=79) |
| Positive | Negative | Uninfected and vaccinated (n=181) |
| Negative | Negative | Uninfected and unvaccinated (n=655) |
| Negative | Positive | False negative on Vitros and/or false positive on Roche (n=3; removed) |
| Positive | No result | (n=15; removed) |
| Negative | No result | (n=43; removed) |
| No result | Positive | (n=2; removed) |
| No result | Negative | (n=21; removed) |

**Supplementary Table 2:** Bivariate posteriors by ZIP code.

| zcta | Mean,<br>P(vaccin<br>ation) | Mean,<br>P(infecti<br>on) | 2.5%,<br>P(vaccin<br>ation) | 2.5%,<br>P(infecti<br>on) | 50%,<br>P(vaccin<br>ation) | 50%,<br>P(infecti<br>on) | 97.5%,<br>P(vaccin<br>ation) | 97.5%,<br>P(infecti<br>on) | n_sampl<br>es |
| --- | --- | --- | --- | --- | --- | --- | --- | --- | --- |
| 94102 | 0.166 | 0.124 | 0.073 | 0.048 | 0.164 | 0.121 | 0.273 | 0.224 | 36 |
| 94103 | 0.097 | 0.169 | 0.025 | 0.079 | 0.091 | 0.166 | 0.203 | 0.278 | 31 |
| 94105 | 0.060 | 0.166 | 0.002 | 0.044 | 0.046 | 0.158 | 0.193 | 0.333 | 13 |
| 94107 | 0.156 | 0.059 | 0.066 | 0.008 | 0.152 | 0.053 | 0.266 | 0.147 | 28 |
| 94108 | 0.180 | 0.057 | 0.056 | 0.001 | 0.171 | 0.044 | 0.349 | 0.186 | 12 |
| 94109 | 0.257 | 0.101 | 0.178 | 0.046 | 0.256 | 0.099 | 0.341 | 0.170 | 65 |
| 94110 | 0.198 | 0.178 | 0.123 | 0.108 | 0.198 | 0.176 | 0.279 | 0.256 | 75 |
| 94112 | 0.219 | 0.145 | 0.145 | 0.087 | 0.218 | 0.143 | 0.298 | 0.212 | 86 |
| 94114 | 0.198 | 0.070 | 0.105 | 0.017 | 0.195 | 0.065 | 0.307 | 0.153 | 38 |
| 94115 | 0.359 | 0.023 | 0.258 | 0.001 | 0.357 | 0.016 | 0.467 | 0.080 | 39 |
| 94116 | 0.135 | 0.056 | 0.066 | 0.014 | 0.132 | 0.051 | 0.219 | 0.120 | 48 |
| 94117 | 0.177 | 0.042 | 0.097 | 0.006 | 0.175 | 0.037 | 0.273 | 0.108 | 41 |
| 94118 | 0.217 | 0.041 | 0.131 | 0.005 | 0.215 | 0.036 | 0.313 | 0.107 | 43 |
| 94121 | 0.315 | 0.052 | 0.229 | 0.012 | 0.313 | 0.048 | 0.407 | 0.115 | 53 |
| 94122 | 0.292 | 0.058 | 0.210 | 0.017 | 0.291 | 0.055 | 0.382 | 0.118 | 65 |
| 94123 | 0.197 | 0.038 | 0.086 | 0.001 | 0.192 | 0.028 | 0.331 | 0.126 | 21 |
| 94124 | 0.075 | 0.155 | 0.019 | 0.077 | 0.070 | 0.153 | 0.163 | 0.254 | 40 |
| 94127 | 0.322 | 0.097 | 0.166 | 0.015 | 0.317 | 0.088 | 0.494 | 0.235 | 17 |
| 94131 | 0.357 | 0.031 | 0.232 | 0.001 | 0.355 | 0.022 | 0.491 | 0.111 | 28 |
| 94132 | 0.299 | 0.162 | 0.185 | 0.073 | 0.298 | 0.159 | 0.421 | 0.266 | 40 |
| 94133 | 0.215 | 0.067 | 0.106 | 0.011 | 0.211 | 0.060 | 0.348 | 0.161 | 25 |
| 94134 | 0.257 | 0.231 | 0.136 | 0.130 | 0.255 | 0.227 | 0.392 | 0.347 | 36 |

**Supplementary Table 3:**
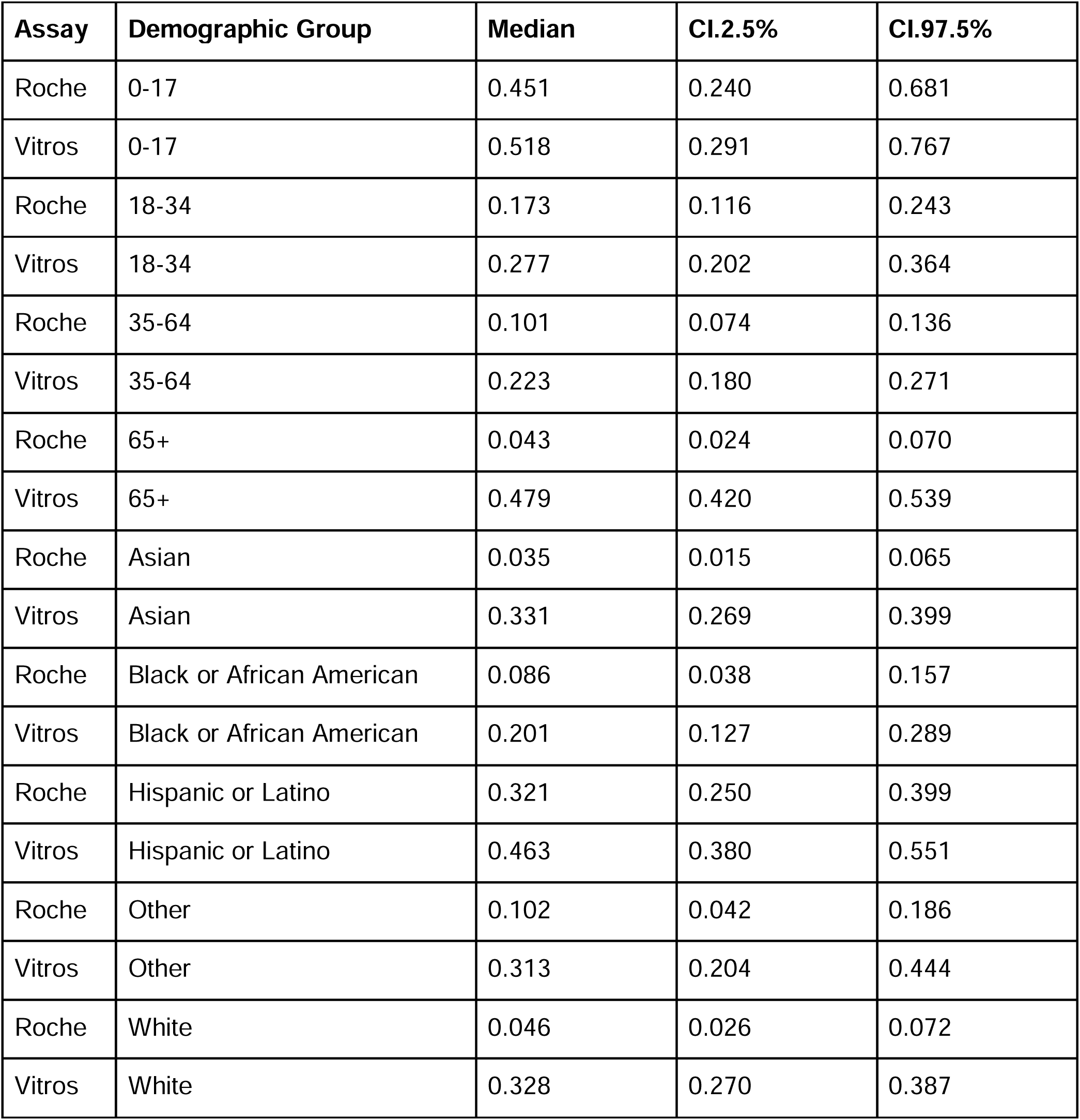
Univariate posteriors by demography.

**Supplementary Table 4:**
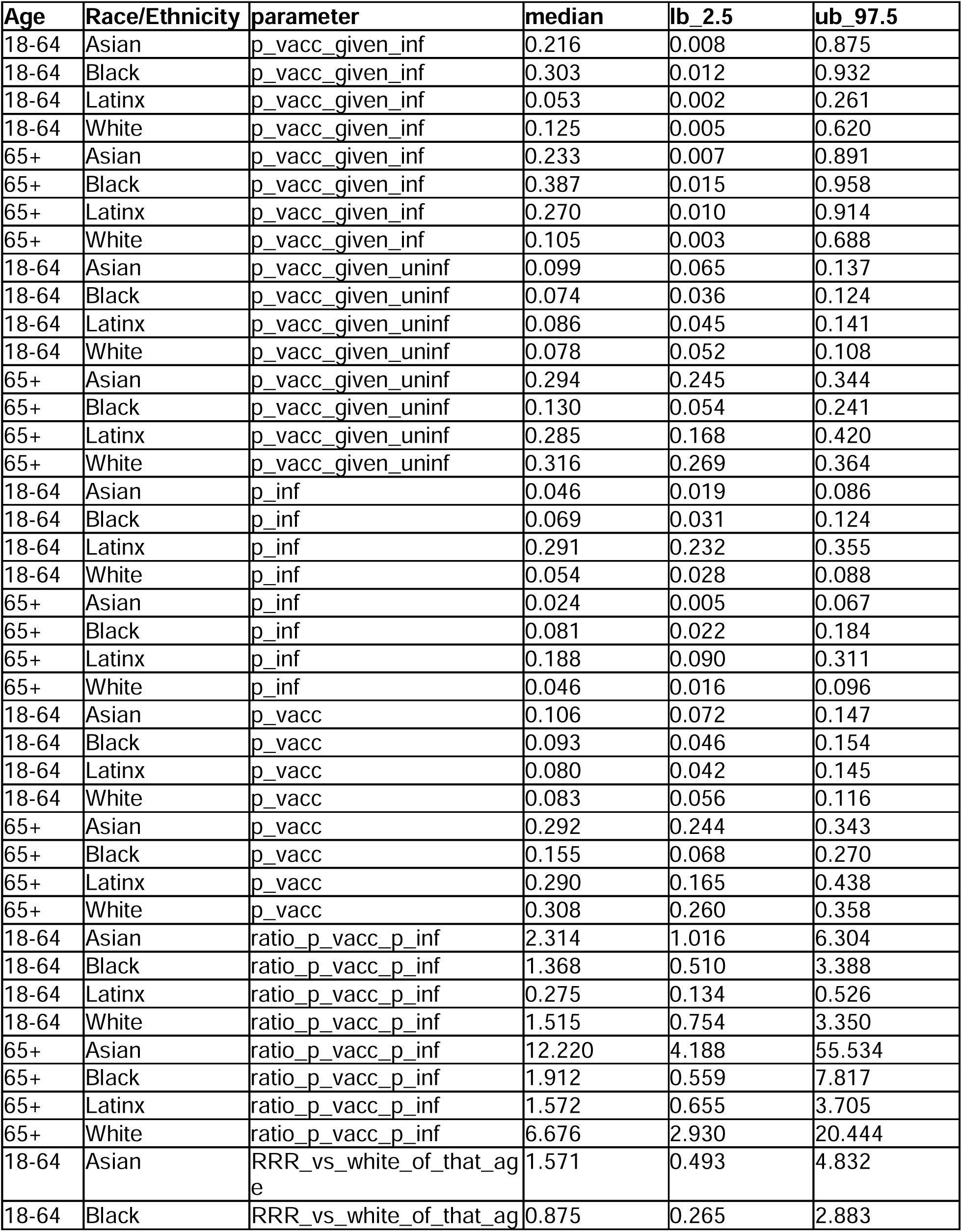
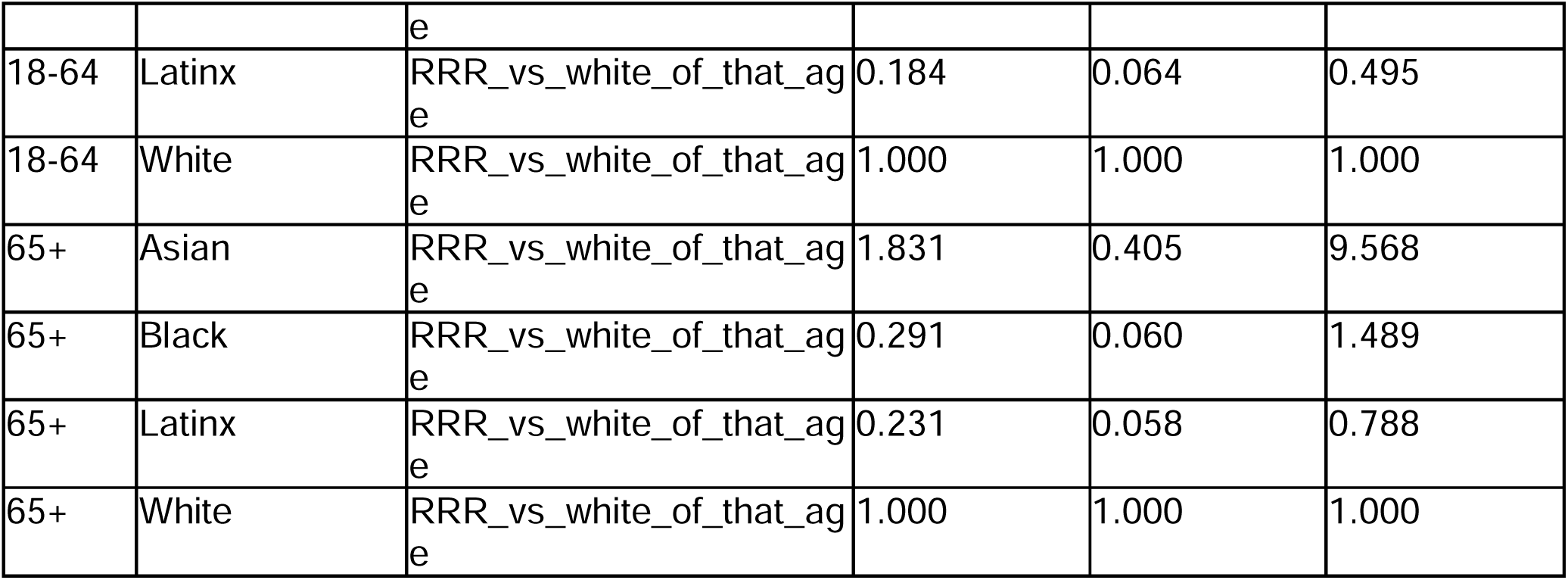
Bivariate antibody results by age and race/ethnicity. Probability of vaccination given infected, probability of vaccination given uninfected, probability of infection, probability of vaccination, probability of vaccination divided by probability of infection (“risk ratio”), and relative risk ratio with a reference group of White race/ethnicity and the corresponding age.

## Data sharing statement

All analysis was conducted using the R statistical software and the Stan programming language. All code to reproduce these results are available at: https://github.com/EPPIcenter/scale-it-2.

## Acknowledgements

We acknowledge sources of funding support, including from the Schmidt Science Fellows, in partnership with the Rhodes Trust (ST); Chan Zuckerberg Biohub Investigator program (BG); the ZSFG Department of Medicine and Division of HIV, ID, and Global Medicine; the MIDAS Coordination Center COVID-19 Urgent Grant Program (MIDASNI2020-5) by a grant from the National Institute of General Medical Science (3U24GM132013-02S2) (IR, ST, IRB); and the National Institutes of Health/National Institute of General Medical Sciences R35GM138361-02 (IRB). We acknowledge Dr. Carina Marquez for helpful comments on the manuscript. We acknowledge Valerie Green and Phillip Williamson at Creative Testing Solutions for performing the Roche testing. We acknowledge the groups of Dr. Kara Lynch and Dr. Alan Wu for facilitating the collection of samples at ZSFG. We acknowledge the groups of Dr. Lee Besana and Dr. Marcelina Coh for facilitating the collection of samples at UCSF.

## Research in context

### Evidence before this study

We searched PubMed on October 13, 2021, using the following search terms: (inequalit* OR disparit*) AND (vaccination OR vaccine) AND (infect* OR exposure) AND (COVID* OR SARS-CoV-2 OR SARS-CoV), published in 2021 (i.e. after the roll-out of vaccination) with no language restrictions. We identified 145 publications, including original research, commentaries, systematic reviews and meta-analyses and reviews. Although many studies discussed or quantified disparities in either vaccination or infection risk, we did not find any studies which measured the combined effect or interaction between disparities in vaccination and infection risk, or quantified the effect of these disparities on population-level immunity.

### Added value of this study

To our knowledge, this is the first study to jointly measure disparities in vaccination and infection using serological data. Serosurveillance systems leveraging demographically-stratified residual serum samples, like the one we implemented, are affordable and flexible, and could be used by Public Health systems to monitor health disparities as they avoid some of the biases inherent in metrics such as case counts and test positivity rates. We also show how vaccination coverage may be monitored, which can be challenging when integrating data from multiple healthcare systems and when denominator populations may be inaccurate for calculating vaccination coverage.

